# Baseline expression levels of drug metabolizing enzymes in non-small cell lung cancer biopsies show promising biomarker and target potential for long-term response to pembrolizumab

**DOI:** 10.1101/2025.11.10.25339842

**Authors:** Kirsten De Ridder, Chiara Maria Vinchesi, Philippe Giron, Nazari Sohrab, Kristof Cuppens, Korkmaz Elif, Ezgi Ulas, Idris Bahce, Lore Decoster, Cleo Goyvaerts

## Abstract

Long-term responders (LTRs) to immune checkpoint inhibitor (ICI) therapy in advanced non-small cell lung cancer (NSCLC) are rare. As no robust biomarkers for LTRs have been defined so far, oncologists are unable to define the optimal treatment duration for this cohort. Hence, standard practice of ICI administration for at least 2 years, regardless of response, does not rely on a solid biological rationale. In this multi-center retrospective study, we analyzed baseline clinical, genetic, and transcriptomic profiles of 14 non-responders (NRs) and 21 LTRs, defined by disease progression shortly after treatment initiation and no evident progression for at least one year after ICI cessation, resp. Comparative analysis of clinical and serological parameters identified an adenocarcinoma histology, low neutrophil-to-lymphocyte ratio (NLR) and low platelet-to-lymphocyte ratio (PLR) to be significantly associated with LTRs. Genetic profiling demonstrated that LTRs were twice as frequently associated with mutations in *KRAS* (56.3% in LTRs, 25% in NRs) while the opposite held true for mutations in *STK11* (25% in NRs, 12.5% in LTRs). Transcriptomic analysis of tumor biopsies from 5 LTRs and 5 NRs treated with pembrolizumab monotherapy at the Universitair Ziekenhuis Brussel (UZB, Belgium) revealed that LTRs show a significantly raised adaptive immune profile, predominantly characterized by T cell- and B cell-related immunity. In addition, we found LTRs to have a significantly lower expression of a drug metabolizing enzyme (DME) gene signature, mainly defined by isotypes of the uridine diphosphate glucuronosyltransferase 1A (*UGT1A*) gene family. The latter was validated in additional biopsies from UZB, Jessa Ziekenhuis (Hasselt, Belgium), and Amsterdam Universitair Medisch Centrum (The Netherlands), as well as in a publicly available dataset. Functional validation using an *in vitro* human tumor cell specific T cell killing assay demonstrated that diclofenac and atazanavir, inhibitors of the DME’s UGT1A enzyme family, enhance ICI efficacy. These findings suggest that baseline expression of the DME signature may inform personalized ICI treatment strategies and support further investigation into the co-administration of ICIs with clinically approved DME inhibitors to improve long-term response rates in advanced NSCLC.

**Graphical Abstract:** 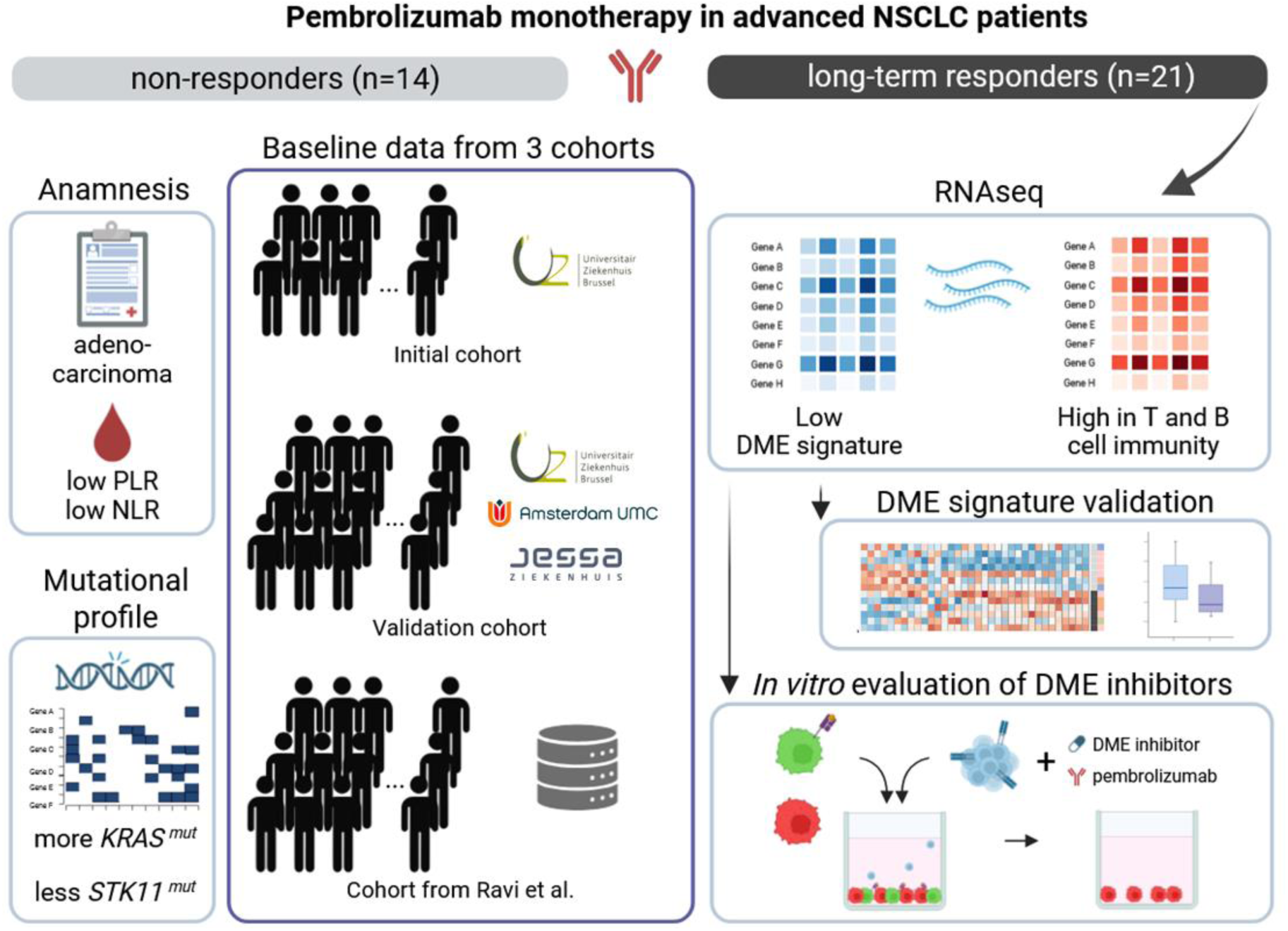

## Introduction

With its discovery thirty-three years ago, the Programmed Death protein 1 (PD-1) and its ligand PD-L1 have revolutionized the therapeutic landscape of lung cancer (Ishida et al., 1992). Especially for advanced non-oncogene driven non-small cell lung cancer (NSCLC) patients, immune checkpoint inhibitors (ICIs) against the PD-1 pathway became a cornerstone of their first line treatment regimen (Hendriks et al., 2023; Singh et al., 2022). Notably, immunotherapy is the only treatment on the market with the potential to result in long-term response (LTR), lasting for many years after ICI discontinuation in patients with advanced NSCLC. Within a large, multi-center, real-world cohort of 1,050 advanced NSCLC patients first-line treated with pembrolizumab (anti-PD-1) monotherapy, an objective response rate of 49.6% was reported of which 32 patients or 3% showed a durable complete response (CR) (Cortellini et al., 2025). In a meta-analysis study including 4,803 NSCLC patients from nine randomized controlled trials, the incidence of CRs in NSCLC patients treated with ICIs was 1.5% compared to 0.7% in the chemotherapy cohort (J. Li et al., 2019). In a retrospective analysis of 361 patients with advanced solid tumors (of which 80% NSCLC) receiving ICI monotherapy or with concomitant chemotherapy, even up to 11.3% demonstrated a CR (Catozzi et al., 2024).

Although promising, these numbers indicate that only half of patients respond and that within the responders the majority relapses over time leaving a sobering 1.5-11% to be long-term responders (LTRs). Irrespective of these numbers, current standard practice limits ICI administration to 2 years (Hendriks et al., 2023). Ideally, duration of treatment should be adjusted to clinical efficacy and tolerability, especially for LTRs where management optimization could result in shortened and reiterated ICI cycles. This could drastically reduce health care costs, while potentially lowering installation of acquired resistance and onset of serious immune related adverse events (irAEs) (Chan, 2017; Marron et al., 2021; Putzu et al., 2023). Hence reliable biomarkers that can specifically predict LTRs could foster economic and patient-tailored treatment optimization.

The histological tumor proportion score (TPS) of PD-L1 emerged as the only globally approved predictive biomarker for response to ICI in NSCLC (Vitale et al., 2025). Nevertheless, several phase III studies have been unable to reproduce a robust association between PD-L1 expression and ICI effectiveness, reflecting the inaccuracy of the TPS in terms of predictive value (Fitzsimmons et al., 2023). Other potential baseline forecasters for response to ICI include somatic tumor mutational burden (TMB), microsatellite instability (MSI) and neoantigen variety; tumor immune cell infiltration with focus on CD8^+^ tumor-infiltrating lymphocytes (TILs) and B-cell rich tertiary lymphoid structures (TLS); specific mutations (e.g. *KRAS* versus *STK11*) next to epigenetic and transcriptional signatures. Promising circulating biomarkers include peripheral immune cells such as the neutrophil-to-lymphocyte ratio (NLR), platelet-to-lymphocyte ratio (PLR) and LIPI score (based on the derived neutrophil-to-lymphocyte ratio (dNLR) and lactate dehydrogenase). Furthermore, circulating cytokines, nutritional and inflammation related markers (e.g. C-reactive protein (CRP)/ albumin ratio), circulating tumor cells (CTCs), circulating tumor DNA (ctDNA), exosomes and non-coding RNAs have shown predictive power for ICI efficacy. Additionally, pretreatment history, clinical stage and metastases, gender, body mass index, human leukocyte antigen class I evolutionary divergence and diversity, smoking status and the microbiome have been associated to ICI response (Baseri et al., 2019; Chowell et al., 2019; Cuppens et al., 2022; Goh et al., 2023; Kang et al., 2025; Mino-Kenudson et al., 2022; Ouyang et al., 2025; Rother et al., 2024; J. Zhang et al., 2025). None of these biomarkers however seem to have universal indisputable predictive value. Notably, multimodality biomarker strategies have been associated with improved performance over singular use of PD-L1 TPS, TMB or a transcriptome signature, advocating for the implementation of a multimodal approach in clinical practice (Cristescu et al., 2018; Lu et al., 2019).

Although ample studies report on potential predictive biomarkers for ICI, most compare non-responders (NRs) with responders without distinguishing partial (PR) from CRs and often lacking long-term follow-up making it impossible to define LTRs. This is rationalized by the fact that case reports of exceptional LTRs and CRs to ICI in NSCLC are rare and possibly underreported (Baseri et al., 2019). This lacune fortifies the need for additional molecular, genomic and transcriptomic markers that specifically distinguish LTRs from PRs and NRs to be used in combination with the PD-L1 TPS to better guide treatment decision making (Putzu et al., 2023).

Hence, in this retrospective multi-center study we aimed to identify a hallmark baseline LTR signature based on clinical, genomic and transcriptomic baseline data from advanced NSCLC patients defined by no evident progression determined by RECIST 1.1 measurements on CT scan for at least one year after stopping pembrolizumab electively. NRs in this study are at the other end of the response spectrum marked by disease progression at six weeks to twelve weeks after treatment initiation. In total 21 LTR and 14 NR patients who received pembrolizumab between 2017 and 2024 were included to define and validate a LTR-related signature.

## Methods

### Patients

In this retrospective multi-center study, a total of 35 advanced NSCLC patients treated with pembrolizumab monotherapy as first or second-line treatment were included. Biopsies from 13 advanced NSCLC patients treated at the Universitair Ziekenhuis Brussel (UZB, Belgium) between 2017 and 2021 were described as our ‘initial cohort’. While later this was expanded to a multi-center study including 22 biopsies and clinicopathological data from patients treated at UZB (n=4), Jessa Ziekenhuis (Hasselt, Belgium, n=5), and Amsterdam UMC (The Netherlands, n=11). More details can be found in **Supplementary Table 1**. Patients were divided according to their response. A LTR was defined as no evident progression determined by RECIST 1.1 measurements on CT scan for at least one year after stopping pembrolizumab treatment either electively or because of irAE. NRs by RECIST 1.1 measurements on CT scan at six weeks to twelve weeks after treatment initiation. Their pre-treatment clinicopathological and serological parameters were collected and managed using REDCap electronic data capture tools hosted at UZB. The study was approved by the Medical Ethics Committee of UZ Brussels/VUB with EC number EC-2021-086.

### RNA and DNA isolation

Diagnostic FFPE tumor biopsies were sliced in two x two 10μm slices for RNA and DNA isolation, respectively, using a RM2245 rotation microtome (Leica). Subsequently, RNA and DNA were extracted separately using the Maxwell RSC RNA or DNA FFPE Kit (Promega), respectively, according to the manufacturer’s instructions on the Maxwell RSC 48 Instrument (Promega). Concentrations were determined with the Qubit RNA or dsDNA HS Assay Kit (respectively) and Qubit 3.0 Fluorometer (Thermo Fisher Scientific). Quality control was performed by evaluating fragment sizes with the Fragment Analyzer (Advanced Analytical).

### DNA sequencing

Version 3 of the Solid and Hematological Tumors Gene panel was used for DNA sequencing, consisting of the coding regions and 11 basepair flanked intronic sequences of 380 disease related genes (**Supplementary Table 3**). A total of 150 ng was used for fragmentation and DNA library preparation using the KAPA Hyper Plus Prep Kit (Roche). DNA libraries are captured with the help of the HyperChoice enrichment probes and KAPA Hyper Capture Beads & Reagents (Roche). Captured fragments were amplified and sequenced by 2 x 100 bp paired-end sequencing on the NovaSeq 6000 Sequencing System (Illumina).

Sequenced reads were demultiplexed and subsequently went through a quality control using FastQC (v0.10.0). Reads were aligned to the human reference genome hg19 (ucsc.hg19.fasta) using Burrows-Wheeler Aligner algorithm BWA-mem (v0.7.10). Aligned reads were sorted and checked for quality using samtools (v0.1.19). Duplicated reads were marked with Picards (v1.97). Further optimization and quality control were done using the Genome Analysis Toolkit GATK (v3.3) and Picard, respectively. Coverage in target regions was measured with samtools (v0.1.19) and an in-house developed Rscript (R v4.1.3.). Variant calling of the reads was carried out by (GATK) Mutect2 and annotated using the Alamut batch pipeline v1.4.

### RNA sequencing

An input of 150 ng of total FFPE RNA was used with KAPA RNA HyperPrep Kit, including RiboErase (Roche). Libraries were created according to the manufacturer’s recommendations and amplified with PCR using KAPA HiFi HotStart ReadyMix and Library Amplification Primer Mix (Roche). The amplified libraries were then attached to complementary flow cells and sequenced on the NovaSeq 6000 Sequencing System (Illumina) using a paired-end sequencing (2 x 100 bp) protocol as this facilitates transcriptome analysis, which generated 25 million reads per sample.

After quality controls of all sequenced samples using FastQC, RNAseq reads were assembled and mapped to the human reference genome hg19 using the alignment software STAR. After alignment and mapping, gene-level quantification was performed using HTSeq-count, delivering counts per gene for each sample.

### Differential gene expression analysis

Data analysis was performed using R v4.4.1. Raw data counts were normalized and used for differential gene expression analysis via the DESeq2 package in R (Love et al., 2014). For data exploration, PCA was performed using variance-stabilized counts. Differentially expressed genes were measured comparing biopsies arising from the LTRs to the NR category. Genes were marked as differentially expressed when adjusted p-value < 0.05 and shrunken log2 fold change threshold was set above or below 2 and −2. Plots were generated using the ggplot2 and pheatmap packages in R.

### Gene set enrichment analysis

GSEA on the MSigDB Gene Ontology biological processes and KEGG gene sets was performed using the full list of genes ranked by their log2 fold change between LTRs and NRs via the ’fgsea’ R package. Gene sets were summarized in bar plots each constructed with a cut-off described in the figure legends. Heatmaps linking differentially expressed (DE) genes to their respective gene sets were constructed using the ComplexHeatmap package in R. Gene set variation analysis (GSVA) was performed to obtain an enrichment score per gene signature for each biopsy. A list of all tested gene signatures can be found in **Supplementary Table 2**. Associations between GSVA scores and response cohorts were assessed using linear models implemented in the limma package. Empirical Bayes was applied to obtain t-scores and corresponding p-values. P-values were adjusted for multiple testing using the Benjamini–Hochberg method.

### Cell-type deconvolution

Cell-type deconvolution was performed on all lung tissue biopsies to estimate the cell population indicative for therapy response. Gene expression deconvolution was done using the dampened weighted least squares algorithm provided by the omnideconv package (Sturm et al., 2020; Tsoucas et al., 2019). As a signature matrix, an online available scRNAseq dataset arising from four NSCLC patients was downloaded from the TISCH website (dataset name: NSCLC_GSE117570) (Sun et al., 2021). Annotated cell populations were described as published by Lambrechts et al. (Lambrechts et al., 2018). To manage memory usage of the algorithm, each annotated cell population was trimmed for 1000 cells maximum.

### TCGA mining

Gene expression data from lung adenocarcinoma (LUAD) and squamous cell carcinoma (LUSC) the cancer genome atlas (TCGA) cohorts were downloaded using the TCGAbiolinks package in R. Both ‘primary tumor’ and ‘solid tissue normal’ sample types were selected in the sample.type parameter. Parameters workflow.type = “STAR – Counts” and data.type = “Gene expression quantification” were chosen. Subsequent count normalization and data filtering were performed using the provided packages. Differential gene expression was performed via the TCGAanalyze_DEA function, comparing tumor tissue to normal tissue and selecting DE genes with an adjusted p-value < 0.05 and |log2 fold change| ≥ 2.

### Genomic and transcriptomic analysis of an online NSCLC cohort

The study of Ravi et al., comparing response rates to anti-PD-(L)1 therapy in NSCLC patients focusing on pre-treatment parameters, was used as a validation cohort (Ravi et al., 2023). Clinical annotations, gene mutation profiles, and differential gene expression data were used. The dataset with an initial size of 393 NSCLC patients was filtered for NSCLC patients that were treated with an anti-PD-1 agent and smokers only. Patients were categorized for their best overall response rates into four categories: CR, PR, stable disease (SD), and progressive disease (PD). The two extremes of the response spectrum were chosen to compare genetic profiles to our results. GSVA was performed as described above.

### Target cell-specific killing assay

Target cell-specific killing assays were conducted in a 96-well plate format. The assay is initiated by seeding 2 x 10^4^ H1650 cells per well one day prior to the introduction of CD8^+^ T cells. Each well was prepared with half eGFP^+^ HLA-A2^+^ MAGE-A1^+^ H1650 target cells and half Katushka^+^ (Kat^+^) H1650 non-target cells. DME inhibitors atazanavir or diclofenac were added at concentrations of 50µM, 5µM, and 50µM, respectively, four hours post cultivation. The following day, CD8^+^ MicroBeads MACS sorted (MiltenyiBiotec) PBMCs were electroporated with relevant anti-MAGE-A1 T cell receptor (TCR) mRNA to generate effector cytotoxic T lymphocytes (CTLs) specifically recognizing the HLA-A2^+^ MAGE-A1^+^ target cells. Subsequently, Five hours post-electroporation CTLs were added to the H1650 cells at a CTL:target cell ratio of 1:1. In the case of monocytes added to the assay, CD14^+^ MicroBeads MACS sorted (MiltenyiBiotec) PBMCs are added at 3 x 10^4^ per well. Anti-PD-1 mAb or IC mAb (pembrolizumab, Keytruda, and RecombiMAb human IgG4 (S228P), BioXcell) were added at a 5 µg/ml concentration. To determine the exact percentage of killing and T cell activation, 24 hours after adding CTLs flow cytometry analysis was performed. Non-adherent cells were washed off at the end of the killing assay and upon washing with PBS, cells were stained for viable cells using fixable viability dye eFluor506 (65-0866-14, Invitrogen). Cold FACS buffer i.e., PBS containing 1% bovine-serum-albumin and 0.02% sodium azide (Sigma-Aldrich) was used to wash the cells. Next, staining with markers 41BB PE-Cy5 (clone: 4B4-1, BD) and CD8 APC-H7 (clone: SK1, BD) was performed for 20 min at 4°C in FACS buffer. Adherent H1650 cells were harvested via trypsinization at 37°C for 30 minutes. Subsequently, cells were washed with PBS containing 1% BSA and 0.02% sodium azide (Sigma-Aldrich). Fluorescently labeled cells were evaluated on an LSR Fortessa flow cytometer (BD), while analysis was performed using FlowJo_10.5.3 software. Target cell-specific killing was assessed by analyzing the ratio of target (eGFP^+^) to non-target (Kat^+^) cells using flow cytometry. The following equation was used to calculate the percentage:

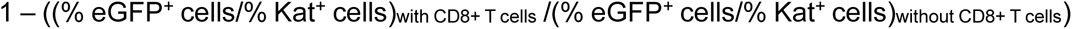

### Statistics

The asterisk number in the figures and tables indicates the level of statistical significance as follows: * for p < 0.05; ** for p < 0.01; *** for p < 0.001 and **** for p < 0.0001. Statistical tests used to determine statistical significance are indicated in the figure legends. Statistical tests were performed using R.

## Results

### Clinicopathologic profiles at baseline show a higher prevalence of adenocarcinoma and significantly lower PLR and NLR values in long-term responders

We retrospectively collected data and biopsies from advanced NSCLC patients treated exclusively with pembrolizumab monotherapy as first- or second-line therapy between 2017 and 2024. Patients for whom only on-treatment data or biopsies were available, as well as non-smokers, were excluded from the cohort. Long-term follow-up allowed stratification into two cohorts: 14 NRs who experienced disease progression shortly after initiating therapy, and 21 LTRs who remained progression-free for at least one year following discontinuation of ICI therapy. Patients’ characteristics and blood parameters were recorded at baseline as summarized in **Table 1**. A Fisher’s exact test showed no significant differences for age and gender between the two response groups. The distribution of histology types differed significantly between LTRs and NRs (P value = 0.0235). Specifically, adenocarcinoma was more prevalent in LTRs (85%) compared to NRs (64.3%), while squamous cell carcinoma showed the opposite distribution. Metastatic disease and a PD-L1 TPS above 50% were observed in the majority of both LTRs and NRs, indicating that these clinical features do not distinguish both groups. As the only two patients found within the PD-L1 TPS <50% were situated in the LTRs cohort, this further highlights the imperfect predictive power of PD-L1 TPS as biomarker for LTRs on pembrolizumab. There was no significant difference between patients receiving immunotherapy as first or second line of treatment. Upon closer examination of treatment history, it was evident that among patients who received pembrolizumab as second-line therapy, treatment response could not be reliably predicted based on the nature or type of their first-line regimen (detailed patient anamnesis, **Supplementary Table 1**). The Eastern Cooperative Oncology Group (ECOG) performance status has been reported as a prognostic factor for NSCLC patients, especially an ECOG score ≥2 significantly reduces the median overall survival (OS) and progression free survival (PFS) (Ahmed et al., 2020; Dall’Olio et al., 2020). Interestingly, in our cohort, two patients with an ECOG score ≥2 achieved a complete response, which contrasts with existing literature.

**Table 1:** Clinicopathologic characteristics at baseline in the NSCLC patients under study. . Patients’ and blood specific parameters have been noted pre-treatment. P-values of numerical variables are calculated via multiple t-tests, corrected for FDR via Benjamini and Hochberg method. P-values of categorical variables are calculated via Fisher’s exact test. Abbreviations: tumor proportion score (TPS), baseline C-reactive protein (CRP), lactate dehydrogenase (LDH), neutrophil-to-lymphocyte ratio (NLR), platelet-to-lymphocyte ratio (PLR), and the derived NLR (dNLR), lung immune prognostic index (LIPI) score to categorizes patients with “poor”, “intermediate” and “good” prognosis depending on the baseline LDH and dNLR.

| Clinical characteristics | Non-responders | Long-term responders | P value |
| --- | --- | --- | --- |
| **Age at diagnosis (NR = 14, LTR = 21)** | 69.1 (56.0 - 86.0) | 67.8 (51 - 83.0) | 0.767 |
| **Gender (NR = 14, LTR = 21)** |  |  | 0.089 |
| Female | 3 (21.4%) | 11 (52.4%) |  |
| Male | 11 (78.6%) | 10 (47.6%) |  |
| **Histology (NR = 14, LTR = 20)** |  |  | **0.0235** |
| Adenocarcinoma | 9 (64.3%) | 17 (85%) |  |
| Squamous cell carcinoma | 4 (28.6%) | 1 (5%) |  |
| Large cell carcinoma | 0 (0%) | 1 (5%) |  |
| Undifferentiated | 0 (0%) | 1 (5%) |  |
| Mixed histology | 1 (7.1%) | 0 (0%) |  |
| **Metastases (NR = 14, LTR = 21)** |  |  | 0.506 |
| Yes | 14 (100%) | 19 (90.5%) |  |
| No | 0 (0%) | 2 (9.5%) |  |
| **PD-L1 TPS (NR = 14, LTR = 21)** |  |  | 0.506 |
| >=50% | 14 (100%) | 19 (90.5%) |  |
| <50% | 0 (0%) | 2 (9.5%) |  |
| **Line of treatment (NR = 14, LTR = 20)** |  |  | 0.267 |
| I | 8 (57.1%) | 9 (45%) |  |
| II | 6 (42.9%) | 11 (55%) |  |
| **Performance status (ECOG) (NR = 14, LTR = 20)** |  |  | 0.286 |
| 0 | 6 (42.9%) | 12 (60%) |  |
| 1 | 8 (57.1%) | 6 (30%) |  |
| 2 | 0 (0%) | 2 (10%) |  |
| **CRP (mg/l) (NR = 8, LTR = 12)** | 66.96 (16.6 - 193) | 20.8 (0.6 – 114.1) | 0.133 |
| **LDH (U/l) (NR = 12, LTR = 14)** | 368.2 (156.0 - 732.0) | 391.5 (128.0 - 654.0) | 0.767 |
| **NLR (NR = 8, LTR = 12)** | 5.9 (3.6 - 10.2) | 3.5 (2.6 – 6.8) | **0.0294** |
| **PLR (NR = 8, LTR = 12)** | 339.8 (221.3 - 450.5) | 185.7 (50.6 – 479.5) | **0.0206** |
| **dNLR (NR = 8, LTR = 12)** | 2.789 (2.1 - 4.9) | 2.1 (1.5 – 3.6) | 0.133 |
| **LIPI score (NR = 6, LTR = 7)** |  |  | 0.380 |
| Intermediate | 6 (100%) | 5 (71.4%) |  |
| Poor | 0 (0%) | 2 (28.6%) |  |

Next, baseline serological values were analyzed to evaluate their potential as liquid biopsy markers for LTR. Neither of the serum inflammation markers CRP and LDH showed a significant correlation with the NR versus LTR group. As the median value of CRP in NRs was 66.96 mg/l compared to 20.8 mg/l in LTRs, the high variability in both groups resulted in an insignificant difference. Also no significances could be noted for the dNLR nor LIPI score. Notably, we can demonstrate that significantly higher NLR and PLR medians were linked to NRs specifically (5.9 versus 3.5 for NLR and 339.8 versus 185.7 in NRs versus LTRs with resp. P values of 0.0294 and 0.0206).

### Long-term responders exhibit a genomic profile characterized by increased KRAS mutations and reduced STK11 alterations

In addition to patient characteristics and baseline serological markers, tumor biopsy samples were examined to investigate the genomic mutational landscape of LTRs and NRs. Although the size of our cohort is limited, aberrations in *KRAS* seemed to be more prevalent in the LTRs (56.3%) compared to the NRs (25%). Additionally, a larger proportion of LTRs had no detectable pathogenic mutations (31.3% and 16.7% resp.), while *STK11* mutations were observed twice as much in NRs (25%) than in LTRs (12.5%) (**Figure 1A**). To fortify our findings, we compared our results with a NSCLC cohort, as published by Ravi et al., 2023. Their initial sample size was 393 NSCLC patients treated with anti-PD-(L)1 therapy. Overall response was divided into PD and SD noted as NRs to therapy, while PR and CR were defined as patients responsive to ICI. By filtering for NSCLC patients that were treated with an anti-PD-1 agent only, removing non-smokers and selecting for the two extremes of the response spectrum (PD versus CRs, defined using RECIST 1.1), we continued with 22 NSCLC patients. Their cohort of six CRs included four patients with *KRAS* mutations while the two patients carrying *STK11* mutations were unresponsive to immunotherapy, which, despite the small sample size, mirrors our observation. Across both our cohort and the Ravi et al. dataset, *TP53* mutations appeared frequently in both response groups without a predictive pattern (**Figure 1B**).

**Figure 1:**
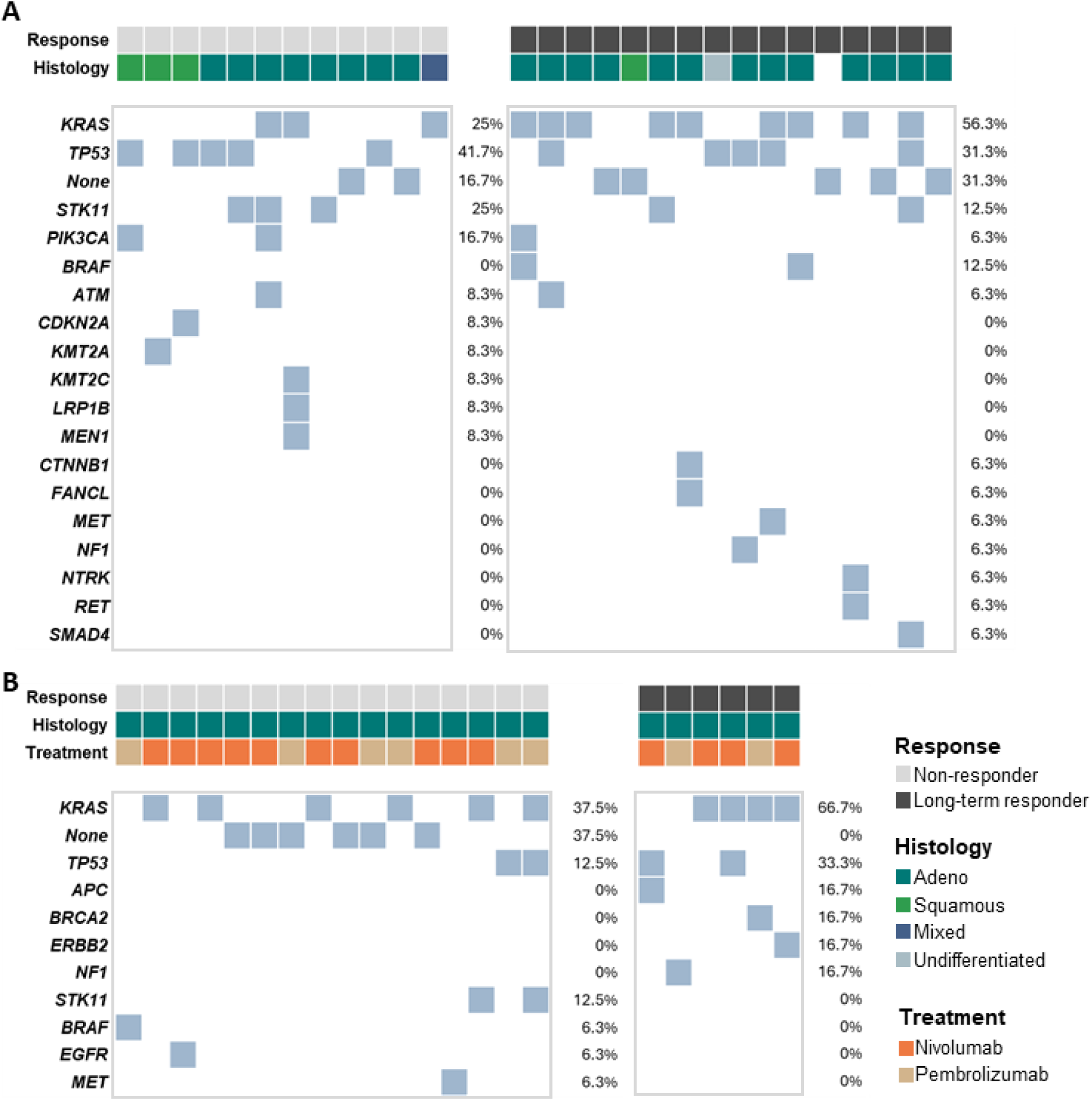
NSCLC patients’ mutational profile in relation to their responsiveness to anti-PD-1 therapy and correlation with publicly available NSCLC data. (**A**) Graphical representation of the pathogenic mutations determined via the ‘solid tumors and hematological tumors gene panel. (LTR n=16, NR n=12). (**B**) Extraction of corresponding mutational profiles of pre-treatment samples from 22 NSCLC patients’ online available data (Ravi et al., 2023). Patients’ responses are categorized into two best overall response groups. NRs are patients categorized as PD (n=16), LTRs are patients noted as CRs (n=6). Percentages in A and B represent the frequency of mutations.

### Validation of the differentially expressed gene profiles of LTRs versus NRs

To elucidate baseline transcriptional expression patterns that could be associated with subsequent response to pembrolizumab, we performed a first bulk RNA sequencing (RNAseq) experiment on curated tumor biopsies from advanced NSCLC patients treated at the UZB (Belgium) between 2017 and 2021 (‘initial cohort’). From the thirteen patients included, nineteen biopsies were sequenced. As five biopsies, originating from liver and brain metastasis, showed expression profiles distinct from all other sampling sites, they were excluded from RNAseq analysis. After QC and filtering this resulted in seven biopsies from five NRs and six biopsies from five LTRs, originating from either primary lung tumor tissue or metastatic lesions (**Supplementary Table 1**).

A principal component analysis (PCA) was performed to observe the variability among samples. First two principal components explained 54% of the overall variance. LTRs clustered together, indicating a distinct transcriptional profile compared to NRs, independent from the tissue origin (**Figure 2A**). Consistently, hierarchical clustering of Euclidean distances showed that biopsies grouped primarily by clinical response, with LTRs forming a separate cluster from NRs. Notably, one LTR clustered closer to the NR cohort, suggesting partial similarity in its expression profile (**Figure 2B**). To identify the differentially expressed (DE) genes between both response categories, a DESeq2 analysis was performed. DE genes were defined with cut-off values |log2 fold change| ≥ 2 and adjusted p-value (adjp) < 0.05. Despite our relatively low sample size, we found 312 up- or down-regulated genes (**Figure 2C**), indicating significant transcriptional baseline differences between both groups.

**Figure 2:**
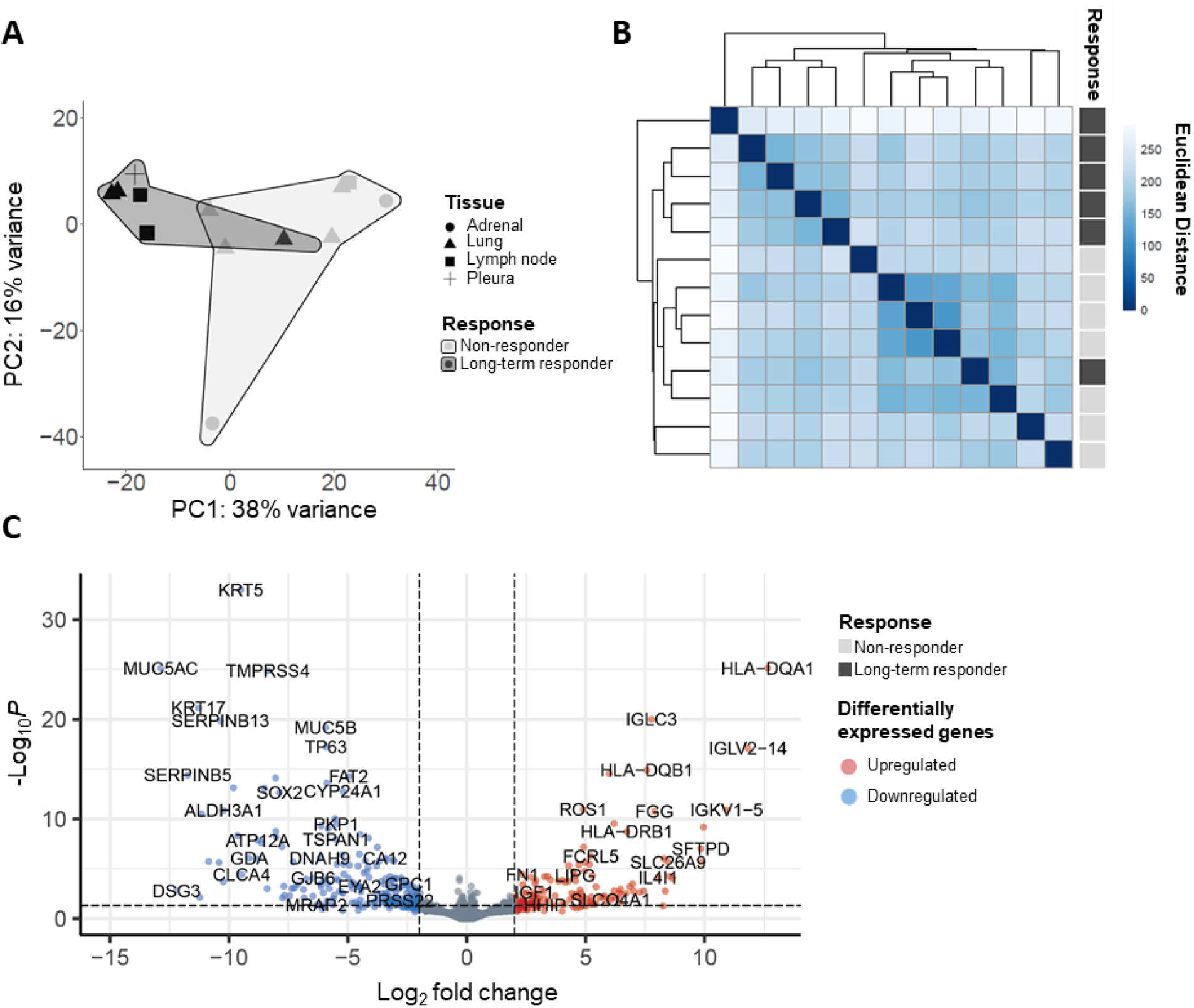
Transcriptomic analysis of pre-treatment multiregional biopsies from long-term responders and non-responders. (**A**) PCA plot representing the first and second principal component (PC) of the 14 samples included in this analysis. The plot depicts the transcriptional profile of both cohorts, clustered by the grey boxes (LTR, n= 6, NR, n=7). (**B**) Distance matrix displaying the sample distribution clustered via Euclidean distance. (**C**) Volcano plot displaying DE genes when adjp < 0.05 and |log2 fold change| > 2. Red dots indicate genes upregulated in LTRs at baseline, while blue dots are highest in NRs pre-treatment.

### High baseline adaptive immune scores correlate with an increased therapy efficacy

To understand which biological processes were significantly more pronounced in the baseline tumor biopsies from LTRs compared to NRs, we performed a gene set enrichment analysis (GSEA). A GSEA analysis on gene ontology (GO) biological processes, using the fgsea package in R, resulted in 30 upregulated pathways (adjp < 0.05) in the LTRs compared to the NRs (**Figure 3A**). As such, four main processes could be described: ‘phagocytosis’, ‘antigen (Ag) processing and presentation’, ‘B cells and immunoglobulin (Ig) production’, and ‘immune response and activation’. The prominence of T and B cell-related pathways highlights a pre-treated LTR-TME defined by an adaptive immunity. With a clear indication for the relevance of immune cells in the pre-treatment TME, we opted for a cell-type deconvolution to estimate which cell populations could be indicative for therapy response. To do so, a publicly available single cell RNAseq (scRNAseq) dataset derived of four NSCLC patients (Lambrechts et al., 2018) was used as a signature matrix. Although none of the differences in immune cell abundances were significant, a trend towards higher baseline levels of monocytes, M2- and especially M1-like macrophages, naive CD4^+^ T cells, CD4^+^ T helper cells and B cells was observed within the LTRs. Observations that are partly in line with the increase in the 4 significantly pronounced processes defined in Figure 3A. Within the non-immune cell fraction, alveolar cells showed higher baseline levels in the LTRs while the opposite held true for malignant cells (**Figure 3B**). The heatmap of all significantly upregulated genes shows a heterogeneous expression pattern across samples, with distinct clusters varying in each response group indicating substantial inter-patient diversity in gene expression profiles (**Supplementary Figure 1**). These features suggest a baseline immune-active state, albeit with considerable patient-to-patient variability.

**Figure 3:**
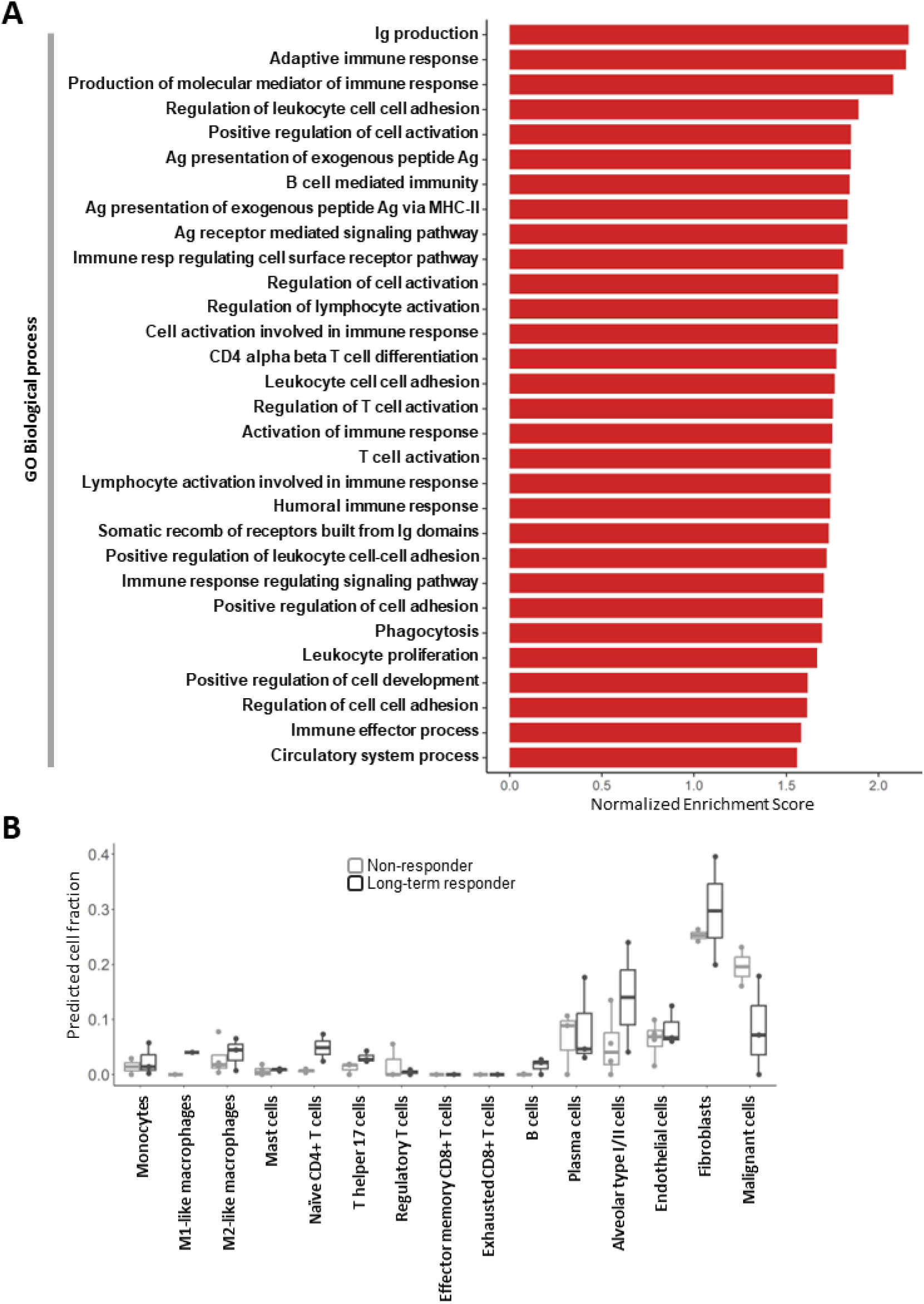
**Long-term responders are marked by higher baseline levels of immune related signatures than non-responders**. (**A**) GSEA results depicted in bar plots representative for the top Gene Ontology (GO) biological processes upregulated in LTRs with an adjp < 0.05. (**B**) Boxplots representing predicted cell fractions calculated via cell-type deconvolution in the lung biopsies (LTR, n=3, NR, n=4). Analysis is based on a signature matrix consisting of 15 cell-type clusters obtained via online available data from Lambrechts et al., 2018. Two-sided Wilcoxon test corrected for FDR via Benjamini and Hochberg method showed no significant differences between LTRs and NRs.

### Subthreshold baseline expression of a drug metabolizing enzyme gene signature is indicative for LTR to pembrolizumab

While upregulated GSEA highlighted the presence of an immune-enriched environment in the LTRs, we also investigated the biological meaning of the downregulated pathways at baseline. GSEA using the KEGG pathway database defined 4 significantly downregulated metabolism related gene sets (**Figure 4A**), predominantly marked by members of the UGT1A gene family (**Figure 4B**). To investigate the expression patterns across all biopsies, we visualized all downregulated DE genes in a heatmap (**Figure 4C**). Hierarchical clustering using Euclidean distance grouped the genes into four major clusters. The largest cluster, marked by the dashed square, is characterized by genes with consistently high expression in the NRs and markedly lower expression in all but one of the LTR biopsies. A GO over-representation analysis on this subset of genes (present in the dashed square), highlights their involvement in three main processes: epidermal cell development, gland development, and xenobiotic metabolic processes (**Figure 4D**). The genes involved in the latter cluster, marked by the dashed circle, are genes known to detoxify exogenous, endogenous and xenobiotics substrates, the so-called drug metabolizing enzyme (DME) genes. LTRs had low baseline expression of three genes from a phase I drug metabolization process, namely cytochrome P450 (CYP) enzymes *CYP19A1*, *CYP2S1*, and *CYP26A1* were downregulated. Moreover, there was a low pre-treatment expression of genes involved in the phase II drug metabolic processes, namely the glucuronidation genes *UGT1A1*, *UGT1A4*, *UGT1A6*, *UGT1A7* from the UDP-glycosyltransferase family as well as *ALDH3A1* from the aldehyde dehydrogenase (ALDH) family as visualized in the heatmap of **Figure 4E**. Exploring gene expression data from TCGA cohorts LUAD and LUSC further confirmed that DME gene expression is significantly higher in tumor tissue compared to adjacent normal tissue (**Supplementary Figure 2**). While DMEs have been linked to therapy resistance in anti-cancer therapy, this is solely known to affect small molecule inhibitors and chemotherapy and has not been linked to ICI-resistance (Heersche et al., 2022; Kaur et al., 2020). Considering the novelty of these findings and the relatively small sample size of our ‘initial cohort’ (5 NRs and 5 LTRs), we selected the defined DME gene set comprising of *ALDH3A1, CYP19A1*, *CYP2S1*, *CYP26A1, UGT1A1*, *UGT1A4*, *UGT1A6*, and *UGT1A7* to further evaluate whether this holds predictive potential as a gene signature to distinguish NRs from LTRs.

**Figure 4:**
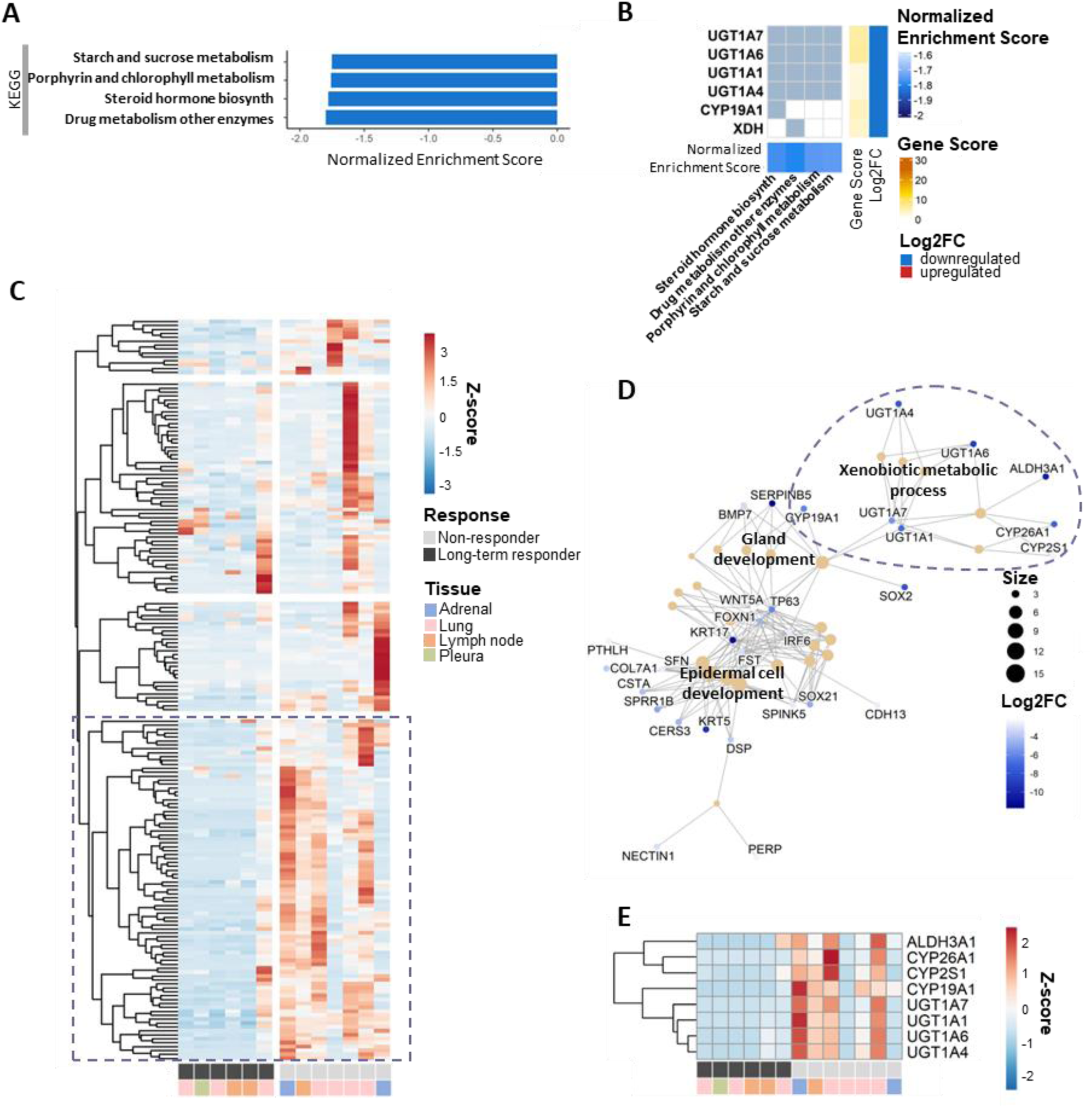
Long-term responders are characterized by the downregulation of pathways involved in xenobiotic and steroid metabolism. (**A**) GSEA results on KEGG pathways depicted in bar plots representing mechanisms downregulated in LTRs. Pathways have a adjp below 0.05. (**B**) Heatmap plot depicts the DE genes involved in the significantly downregulated pathways KEGG pathways. Genes involved in one or more processes are on the y-axis, and the related pathways are displayed on the x-axis. On the right both gene scores (-log10 adjp) and log2 fold changes are shown for the respective genes. Gene sets are depicted with their NES score at the bottom of the graph. (**C**) Heatmap showing the gene expression pattern of all downregulated genes, clustered via Euclidean distance. (**D**) GO over-representation analysis on the largest cluster obtained from (C) depicting the top 25 significantly downregulated biological processes. GO processes are clustered in three main subjects. (**E**) Heatmap visualizing all genes involved in the xenobiotic metabolic processes from (D).

Therefore, we first compiled an overview of publicly available gene sets that were previously associated with ICI response (**Supplementary Table 2**). Throughout a single sample GSEA using gene set variation analysis (GSVA), all signatures, including our DME signature, were given a score to indicate to which response group within our initial cohort each gene signature was most related. Six immune related gene signatures significantly correlated with the LTRs transcriptomic profile, confirming high baseline expression of immune subsets and/or activation. On the other side of the response spectrum, only two signatures were significantly associated with NRs: a MYC-related gene set and the DME signature (**Figure 5A** and **5B**).

**Figure 5:**
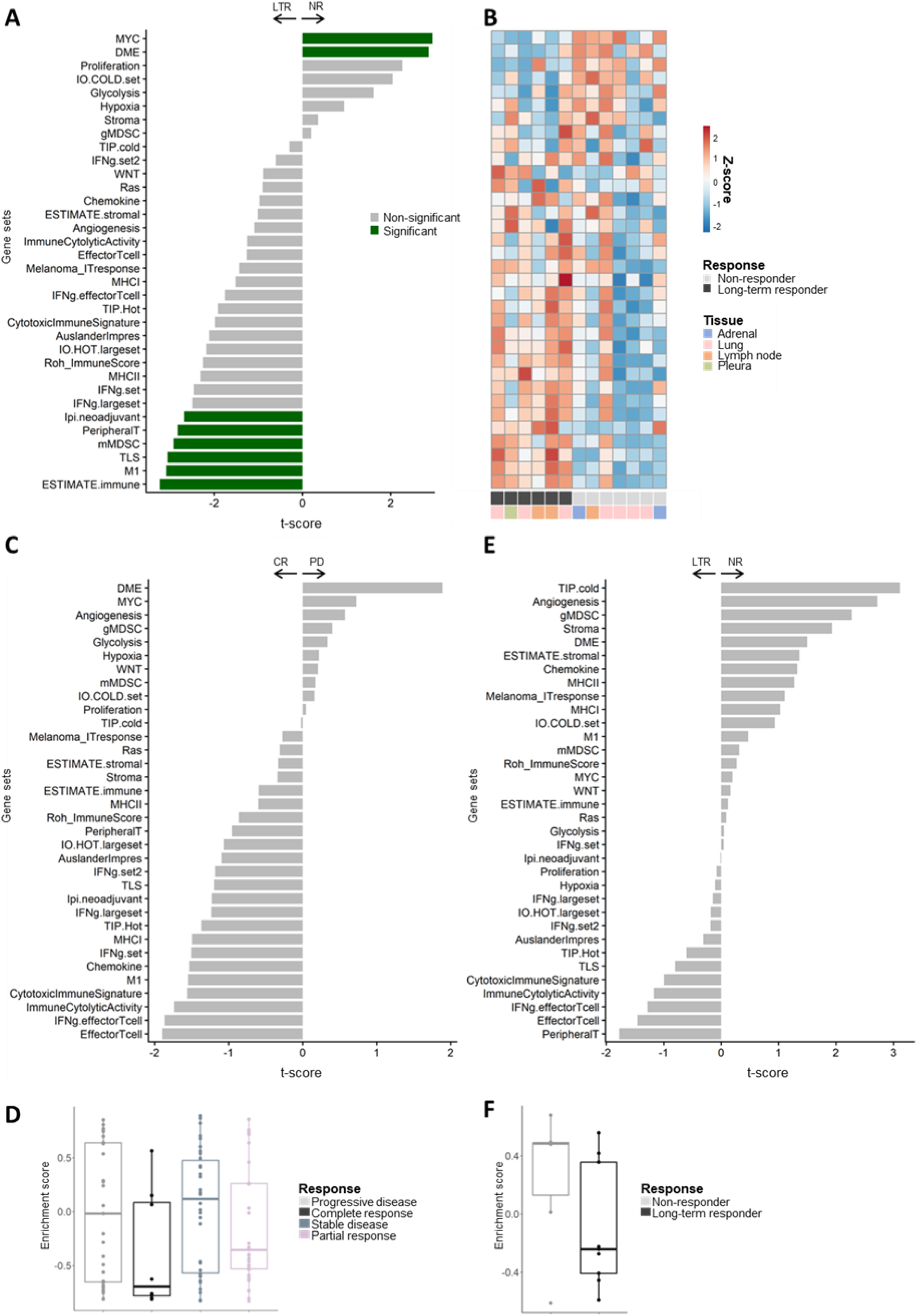
The presence of drug metabolizing enzymes is indicative for poor responsiveness towards anti-PD-1 therapy. (**A**) GSVA between LTRs and NRs performed using 34 gene signatures derived from literature on responsiveness to ICI therapy as well as our own defined DME signature. Significant differences are calculated using the limma package in R. Gene sets with an adjp below 0.05 are considered significant (green). (**B**) Heatmap represents the enrichment scores of each gene set per sample. (**C**) GSVA performed using the 34 signatures on online available data (Ravi et al., 2023). (**D**) Enrichment score calculated for the DME signature displayed for each sample in the publicly available dataset. (**E**) GSVA performed using the 34 signatures on our validation cohort. as well as in the validation cohort (**F**) Enrichment score calculated for the DME signature displayed for each sample in our validation cohort.

Secondly, we interpreted the DME gene set in the context of the online available transcriptomic dataset from Ravi et al., 2023. Within this dataset, we specifically selected the base level normalized counts from the NSCLC patients that showed CR (n=8) or PD (n=31), as a proxy for our LTR and NR cohorts. In contrast to our own RNAseq data from the initial cohort (Figure 5A), comparative GSVA analysis of the counts from the CR versus PD patients of the Ravi et al. dataset, showed no significant correlations with any previously described predictive gene signatures for ICI effectiveness. Notably however, the most predictive score for PD patients in the Ravi et al. dataset was found for the DME signature (**Figure 5C**). **Figure 5D** show the enrichment scores of the DME signature across individual biopsies from all four response groups initially defined in the cohort (n=108). It is clear that the lowest enrichment score is found in the CRs, while increasingly higher when patients show lesser response to ICI therapy.

Thirdly, we created a ‘validation cohort’ by conducting a multi-centric retrospective study with advanced NSCLC patients treated with pembrolizumab at UZB (Belgium), Jessa Ziekenhuis Hasselt (Belgium), and Amsterdam Universitair Medisch Centrum (UMC, The Netherlands). This resulted in the collection and bulk RNAseq analysis of 16 extra tumor biopsies (6 NRs and 10 LTRs). Upon comparative GSVA analysis with previously described predictive gene signatures none showed significant correlation with the NR group in the validation cohort. Nevertheless, the DME signature also showed a correlative trend in the NR group (**Figure 5E**). As such the DME signatures’ correlation with the NR group was found to be consistent for our initial cohort, the dataset from Ravi et al. and our validation cohort. Notably, this consistency did not hold true for most other previously published signatures. In essence the only signatures that were consistently associated with the NR cohorts across all 3 datasets were: DME, gMDSC IO COLD set and Glycolysis, while with the LTRs: IFNg, IO HOT, Immune Cytolytic Activity, Cytotoxic Immune Signature, Effector T cell, Peripheral T cell, IFNg effector T cell, AuslanderImpres, and TLS. **Figure 5F** reflects the lower enrichment scores of the DME signature found in the LTRs compared to NRs across all individual biopsies from our validation cohort.

Overall, we report on a consistent correlation between subthreshold expression of the DME gene set in 3 different LTR cohorts, highlighting its potential to guide treatment design and ultimately improve the likelihood of LTR to pembrolizumab in future NSCLC patients.

### DME inhibition shows potential to revert resistance in an in vitro killing assay

To understand which cell populations drive the aberrant DME expression profile within the TME, we analyzed publicly available scRNAseq data (TISCH2, Lambrechts et al., 2018). This revealed that xenobiotic metabolism linked genes are predominantly expressed by malignant cells and fibroblasts, alongside innate immune cells such as monocytes and M1- and M2-like macrophages (**Supplementary Figure 3**). To further investigate the role of DMEs in anti-PD-1 ICI therapy, a target cell-specific CTL killing assay was designed. In this assay, we tested two clinically available drugs with DME inhibiting properties, namely diclofenac (UGT1A4, UGT1A6, and UGT1A7), and atazanavir (UGT1A1). For this the H1650 eGFP^+^ MAGE-A1^+^ target cell line was mixed and plated with the non-target H1650 Kat^+^ line. Twenty-four hours later, CD8^+^ sorted PBMCs were electroporated with mRNA encoding an anti-MAGE-A1 TCR and added to the H1650 mixture. As monocytes were observed as a potential source of DMEs in the TME, CD14^+^ sorted peripheral blood mononuclear cells (PBMCs) were also included (**Figure 6A**). Another 24 hours later, target cell-specific CTL killing was defined by measurement of the green to red signal via flow cytometry. The presence of diclofenac and atazanavir did significantly enhance killing (**Figure 6B** and **D**). Furthermore, diclofenac and atazanavir (significantly and non-significantly resp.) boosted the percentage of 41bb^+^ CD8^+^ T cells specifically in the conditions where pembrolizumab was added, be it irrespective of monocytes’ presence (**Figure 6C** and **E**). These results highlight that the co-blockade of PD-1 and the UGT1A enzymes represent an avenue that is worthwhile exploring, especially as UGT1A enzyme blocking compounds are already clinically available.

**Figure 6:**
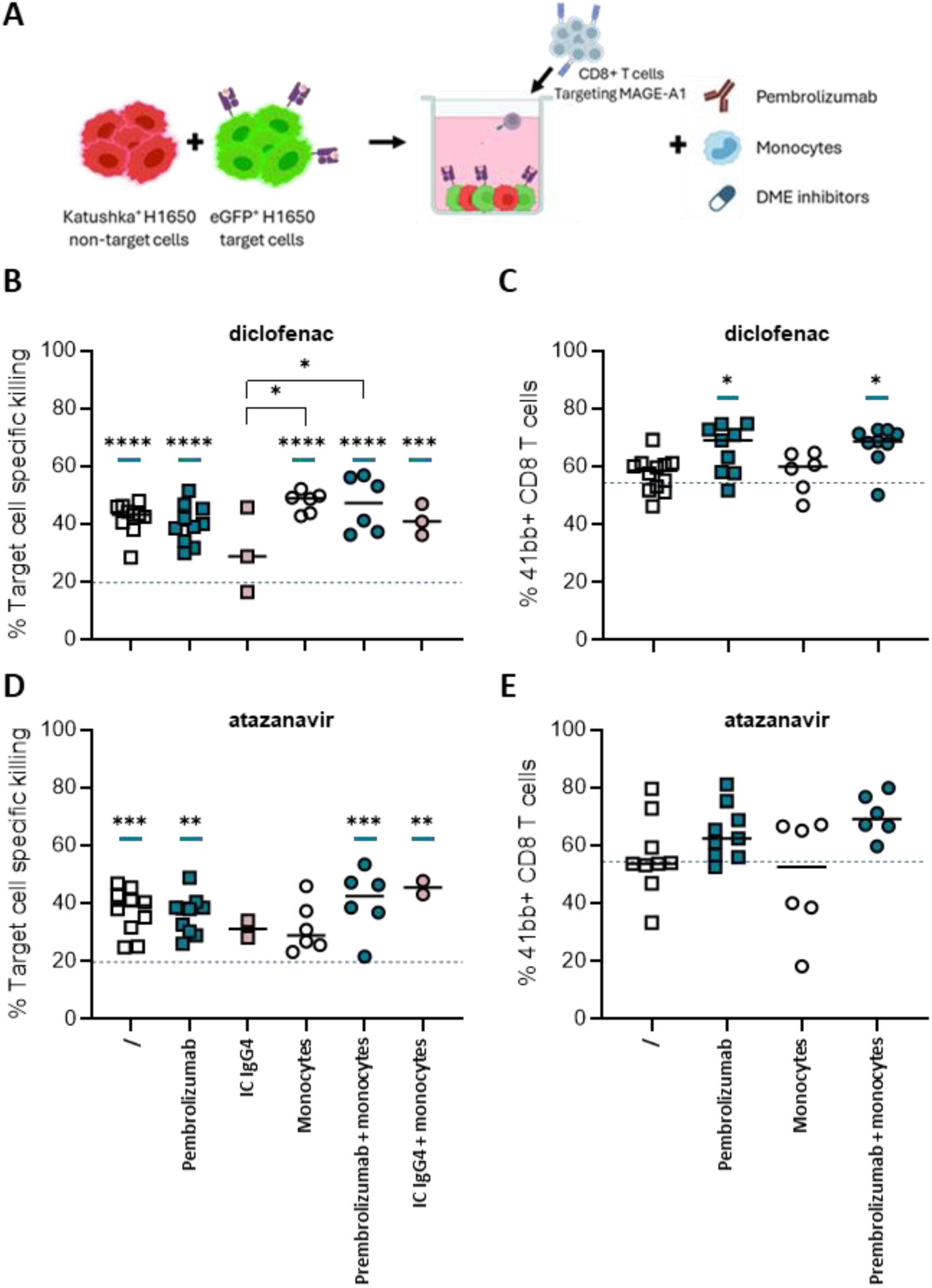
Combination of DME inhibitor with anti-PD-1 mAb pembrolizumab shows potential to boost T cell cytotoxicity. (**A**) Schematic representation of the in vitro target cell-specific killing assay used to investigate the effect of DME inhibitors on ICI efficacy. The MAGE-A1 cancer cell specific killing assay start by co-culturing 10k H1650 eGFP^+^ MAGE-A1^+^ HLA-A2^+^ target cells with 10k H1650 Kat^+^ non-target cells. One day later, MAGE-A1 specific T cells are added at a 1:1 target:T cell ratio. Additionally, 3k CD14^+^ PBMCs are added as a potential source of DMEs. Anti-PD-1 mAb or its IC mAb is added at 5µg/ml per well. (**B**,**D**) Target cell-specific killing percentages were measured one day after T cell addition. The ratio of green to red cancer cells is measured via flow cytometry (n=1-3, 2-3 replicates/exp). The DME inhibitors diclofenac or atazanavir are added at a concentration of 50µM and 5µM respectively. Blue lines represent conditions significantly different from the anti-PD-1 mAb condition without inhibitor, of which the blue dashed line represents the average value. (**C**,**E**) Flow cytometry measurement of 41bb^+^ CD8^+^ T-cells at the endpoint of the killing assay (n=2-4, 3 replicates/exp). Measurements are performed at the end of the killing assay in the presence of DME inhibitors. Blue lines represent conditions significantly different from the anti-PD-1 mAb condition without inhibitor, of which the blue dashed line represents the average value. A one-way ANOVA with Tukey’s multiple comparisons test was performed to determine statistical significance in panels **B**-**E**.

## Discussion

In this retrospective, multi-center study, we compared NSCLC patients categorized in the two extremes of the response spectrum to ICI monotherapy. We aimed to define features with a predictive potential for NRs versus LTRs to aid clinicians during patient-tailored treatment regimen optimization. For this, we analyzed clinicopathologic characteristics, genomic aberrations, and transcriptomic profiles at baseline.

PD-L1 TPS remains the primary criterion for patient stratification. Contra-indicatively, all 14 NRs showed a PD-L1 TPS of ≥ 50% while a <50% TPS was only observed in 2 LTRs. This observation reinforces the notion that the predictive value of the PD-L1 TPS is limited (Fitzsimmons et al., 2023; Mok et al., 2019; Rittmeyer et al., 2017). Our data further demonstrate that the adenocarcinoma subtype has potential predictive value for the LTR cohort, yet this needs further validation. Especially as previous reports on the clinical impact of histological subtype on ICI efficacy have been heterogenous (Carbone et al., 2017; Hellmann et al., 2019; Herbst et al., 2016; F. Li et al., 2022; Mok et al., 2019; Reck et al., 2016). In the evaluation of serological values from both response groups, we could correlate the serological PLR and NLR to therapy outcome. Our findings align with meta-analyses linking high baseline PLR and NLR to inferior OS and PFS in NSCLC and advanced cancers (Su et al., 2025; Tsai et al., 2024; N. Zhang et al., 2020; Zhou et al., 2022). In contrast, we were unable to link any other studied parameter (age, gender, ECOG, CRP, LDH, dNLR nor LIPI score) to LTRs versus NRs. A finding we partly attribute to the limited size of our cohort as well as the fact that we specifically compared LTRs with NRs while most studies included PRs and SD in the responder versus non-responder groups, resp.

Aberrations in oncogenes or tumor suppressor genes has been linked to ICI sensitivity (Kalbasi & Ribas, 2019; Keenan et al., 2019). In line, analysis of our dataset and that of Ravi et al. revealed twice as many LTRs with *KRAS* mutations compared to the NR cohort while *STK11* mutations were detected in the majority of NRs. In accordance, co-mutation of *KRAS* and *TP53* versus *KRAS* and *STK11*, have been associated with increased response versus primary resistance to PD-1 blockade (Skoulidis et al., 2018).

Beyond the role of genomic alterations in therapy outcome, the TME’s transcriptomic landscape has emerged as potential ICI efficacy predictor as well (Coleman et al., 2023). While most predictive gene signatures have concentrated on immune cell abundance or immune-related processes, our findings show limited reproducibility across the 3 cohorts. To illustrate, while the previously defined ESTIMATE immune, M1 and Ipi neoadjuvant signatures are significantly correlated with LTR in the initial cohort, this is not consistent in the other two LTR cohorts. In contrast, the Peripheral T cell and TLS-related signatures showed consistent correlation with LTR in all three cohorts, underscoring the importance of a T-cell rich TME at baseline and the favorable impact of TLS on ICI efficacy. Notably,

GSEA and cell deconvolution showed increase in biological processes related to T and B cell-related immunity within the LTRs of the initial cohort. This fortifies the correlation between LTRs and the Peripheral T cell and TLS-related signatures. In line, a recent study on the clinicopathological and serological values of 42 LTR NSCLC patients treated with chemo-immunotherapy noted a high baseline abundance of circulating B cells (Huang et al., 2025). While the above-mentioned LTR-specific features show encouraging predictive value, prior associations of histology, serological parameters and mutational profile with prognosis suggest that cautious interpretation and further validation are needed. Moreover, future efforts should aim to develop multifaceted signatures rather than relying on a single predictive marker.

We hypothesized that the baseline transcriptomic profile of NRs could reveal mechanisms underlying the lack of an inflamed TME and/or identify targets to overcome primary resistance to ICI therapy in patients. Our analysis highlighted xenobiotic, steroid, and xenobiotic metabolism pathways that include genes from the UGT1A, CYP, and ALDH families encoding for DMEs. Dysregulation of the cancer cell metabolome is a known mechanism promoting tumor growth (Rosario et al., 2018). Intratumoral expression of DMEs and their role in metabolite biotransformation is has been linked to cancer progression across tumor types (Allain et al., 2020; Duguay et al., 2004). In lung cancer, we and others have reported on their expression in LUAD and LUSC tissue compared to healthy lung tissue (Liu et al., 2023). While previous reports highlight their prognostic value, DME’s association with therapy outcome has primarily been attributed to clearance of chemotherapeutics and small-molecule inhibitors, contributing to therapy resistance (Verma et al., 2019). However, DMEs cannot metabolize monoclonal antibodies, suggesting another mechanism underlying the observed correlation between low baseline DME levels and LTR to ICI. A possibility lies in their potential to influence bioactive endogenous metabolites levels, thereby shaping the TME in ways that hinder ICI efficacy. While the mechanism behind this is not described, a previous report in inflammatory-associated disease has linked immunological and metabolic pathways, showing a downregulated DME profile in the presence of inflammatory mediators (Stanke-Labesque et al., 2020).

Despite the limited knowledge about the involvement of DMEs in cancer immunotherapy, we are not the first to note a link between xenobiotic metabolism and cancer ICI. Prior studies in renal cell carcinoma similarly associated baseline expression of xenobiotic metabolism signatures to ICI resistance, more precisely the upregulation of UGT1A genes in the NR group (Ascierto et al., 2016; Au et al., 2021). In colon cancer a link between CYP19A1 and immune evasion was noted. Moreover, its protein expression was correlated to infiltration of macrophages, cancer associated fibroblasts, and endothelial cells (Liu et al., 2023). A recent study into the predictive effect of metabolic pathways in advanced NSCLC treated with ICI identified steroid metabolism as a negative predictor of therapy efficacy (Wang et al., 2026). While baseline samples from LTRs in our validation cohort LTRs tend to show a lower mean score, the considerable variability suggests that larger cohorts are needed to confirm the biomarker potential of this signature.

In our *in vitro* target cell-specific killing assay we tested the target potential of these enzymes, using diclofenac and atazanavir as inhibitors of UGT1A enzymes. Both inhibitors had a profound effect on the killing capacity of CTLs, especially improved in the presence of pembrolizumab. Hence we hypothesize that DMEs’ effect on T cell killing can be found in the tumor’s immunometabolism rather than direct antibody clearance. Potential limitations of this *in vitro* assay include off-target effects of diclofenac and atazanavir, as both drugs have biochemical and pharmacokinetic properties beyond UGT1A inhibition. For instance, diclofenac has been shown to enhance T cell activity by decreasing lactate levels ultimately augmenting the anti-PD-1 mAb effect (Renner et al., 2019). For this, DME inhibition on the RNA level is advisable, e.g. through short interfering RNA’s targeting DME genes of interest. With no literature found on the DME expression by monocytes and our results showing no benefit from monocyte presence, further research is needed to identify the main source of DMEs.

In conclusion, this retrospective multi-center study defined a comprehensive LTR-specific signature, based on serological and transcriptomic features. This signature is characterized by a histological adenocarcinoma profile next to low baseline PLR and NLR scores. Moreover, significantly higher expression levels of genes related to T and B-cell immunity were noted in LTRs compared to NRs. While these findings are in line with previous studies, novel is the finding that the LTR signature is also characterized by low to absent baseline expression of DME genes from the CYP, UGT1A and ALDH family. Our *in vitro* findings further suggest that the UGT1A family represents a promising target to boost anti-cancer T cell killing, warranting its further investigation. Together, this multivariable signature underscores the need for integrated biomarkers to aid in the optimization of patient’s ICI treatment regimen, while highlighting DME inhibition as a potential strategy to overcome primary resistance.

## Data Availability

All data produced in the present study are available upon reasonable request to the authors

